# Experiences of faculty and scientists with disabilities at academic institutions in the United States

**DOI:** 10.1101/2024.02.12.24302692

**Authors:** Franz Castro, Caroline Cerilli, Luanjiao Hu, Lisa Iezzoni, Varshini Varadaraj, Bonnielin K. Swenor

## Abstract

**Introduction:** People with disabilities are underrepresented in higher education, facing systematic obstacles such as inaccessible institutions and difficulties in obtaining accommodations. This qualitative study aims to shed light on barriers to accessibility and disability inclusion in STEM and research institutions through confidential qualitative interviews with disabled faculty and scientists.

**Methods:** We recruited participants (via virtual flyers) working in the United States (U.S.) as research faculty or scientists that applied for grant funding (last five years), and self-identified as having a disability. Interviews (n=35) were conducted via semi-structured one-on-one live interviews or written interviews to accommodate participants’ needs. Data were analyzed by two study members using content analysis to identify themes and codes until saturation was reached.

**Results:** Themes included identity/visibility, career trajectories, accessibility, accommodations, bias, representation, and inclusion. Some participants reported not disclosing their disabilities at work or during hiring processes due to fear of negative perceptions from peers or potential employers. Experiences around stigma and bias were noted both in professional relationships and when interacting with disability service offices, underscoring difficulties and delays in processes to secure accommodations. Respondents highlighted the issues of lack of disability inclusion and low representation of people with disabilities in academia, elevating the importance of self-advocacy, and of role models and mentors in shaping career pathways for future researchers with disabilities.

**Conclusion:** Faculty with disabilities encounter systematic barriers at academic institutions, and lack of acknowledgement and research on these experiences has held back institutional and policy changes. To reduce disparities for disabled faculty, academic leadership must allocate resources to address ableism, create more inclusive environments, and raise standards beyond ADA compliance.

## Introduction

Approximately 27% of all United States (U.S.) adults living in the community have a disability.^1^ However, people with disabilities are underrepresented in higher education, as they comprise 21% of undergraduate students, 14.7% of college graduates, and 9.1% of doctorate recipients.^2-4^ People with disabilities face systematic barriers in higher education, such as inaccessible institutions,^5^ difficulties obtaining accommodations,^6,7^ leading to lower enrollment and graduation rates,^8^ and underrepresentation in the academic workforce.^2^

There has been a recent emphasis on increasing disability inclusion in science, technology, engineering, and mathematics (STEM) education and the scientific workforce,^9^ underscoring the importance of further understanding obstacles for disabled people throughout STEM career pathways. While previous research highlights challenges faced by students with disabilities in higher education,^10,11^ there are few studies examining the experiences of research faculty with disabilities in the U.S.^12^ Among this work, commonly documented issues for faculty with disabilities are navigating disability disclosure in the workplace, obtaining disability accommodations, facing stigma and discrimination from colleagues, and contending with structural ableism embedded within institutional policies.^13^

This study aims to understand barriers to accessibility and disability inclusion in the STEM and research settings through confidential qualitative interviews with disabled faculty and scientists.

## Methods

### Study design and participants

This study was approved by the Johns Hopkins Medicine Institutional Review Board (#00310675). Our research relied on various theoretical frameworks, including human right theories and the social model of disability, acknowledging the role of societal barriers, ableism, and exclusion in shaping career pathways.^14^ In order to collect data on the experiences of faculty and scientists with disabilities in the U.S., we recruited participants via virtual flyers followed by an online screening survey created on Qualtrics, circulated through professional networks and social media. Recruitment materials were developed to be compatible with screen reader software and included image descriptions, high-contrast, and alternative texts. The screening survey, open from March to May 2022, assessed whether participants met inclusion criteria for the study: having worked in the U.S. as research faculty or scientists, having applied for research grant funding in the past five years, and self-identifying as having one or multiple disabilities. Disability was defined to be inclusive of people who are d/Deaf, blind or have low vision, have mobility (including upper and/or lower limb mobility), learning, or cognitive disabilities, chronic conditions, psychiatric disabilities, or mental illnesses. Exclusion criteria were inability to provide consent, being unable to speak English, and not providing phone or email addresses to being further contacted by researchers. This screening survey also included a brief description of the study, collected demographic data on all respondents, and asked respondents if they preferred to respond via one-on-one live virtual interviews or written interviews. Participants who selected one-on-one interviews were asked to share any accessibility or accommodation requests, such as Communication Access Realtime Translation (CART) captioning or American Sign Language interpretation. All eligible participants were selected and contacted by a researcher for next steps.

### Data collection

Data collection took place between April and July of 2022. The interview guide, used to lead both the semi-structured one-on-one interviews and the written response prompts, included ten open-ended questions based on a literature review and lived experiences of the study team, and two pilot interviews with two research faculty with disabilities were conducted (Supplementary Table 1). All participants provided informed consent prior to either engaging in a one-on-one interview or completing a written interview.

Participants who preferred to respond via one-on-one interviews scheduled with the interviewing researcher over email. The semi-structured one-hour one-on-one interviews were conducted via Zoom by a study team member with a disability and experience in conducting research interviews. To maximize accessibility, access requests were addressed in advance and participants were invited to engage in the way most comfortable to them, such as with their camera on or off or responding to questions verbally or by typing in the chat. Only audio was recorded from these interviews, and recordings were deidentified and then transcribed by a professional service (Landmark Associates, Phoenix, AZ).

For selected participants who preferred to respond via written interviews, a researcher emailed the study interview questions hosted on Qualtrics (Provo, UT). Participants were given the option to type responses or to record voice messages to respond to each question, which were transcribed by researchers.

### Analysis

Two study members analyzed both the one-on-one and written interview transcripts using content analysis. Study team members independently coded each transcript in different documents for emergent themes and codes using an iterative framework until data saturation was achieved. Team members met regularly to produce a codebook and find consensus for each transcript. Disagreements were resolved by revisiting code definitions until an agreement was reached.

Researchers used Microsoft Word to highlight relevant text in the transcripts and typed codes using the comments functionality. The R (v 4.3.1, 2023) package *docxtractr* (v 0.6.5, 2020) was used to extract all codes from individual transcripts into .csv files. We compared these typed codes with the original codebook and found mismatches due to typos, and utilized the R package *stringdist* (v 0.9.10, 2022) to carry out approximate string matching and retrieve the correct codes. The accuracy of the matches was assessed by one of the study members.

## Results

### Study Population

Eighty-seven individuals responded to the screening survey, of which 14 participated in semi-structured one-on-one interviews and 21 provided responses via written interviews, for a total of 35 responses (Figure 1). Table 1 shows respondents’ disability types. To protect participant confidentiality, breakdown of minority reported demographic characteristics are not shared in this paper. Three major themes emerged that captured participants’ experiences: disability identity and bias, accommodations, and disability advocacy. Table 2 show all subthemes identified in the study.

**Figure 1.**
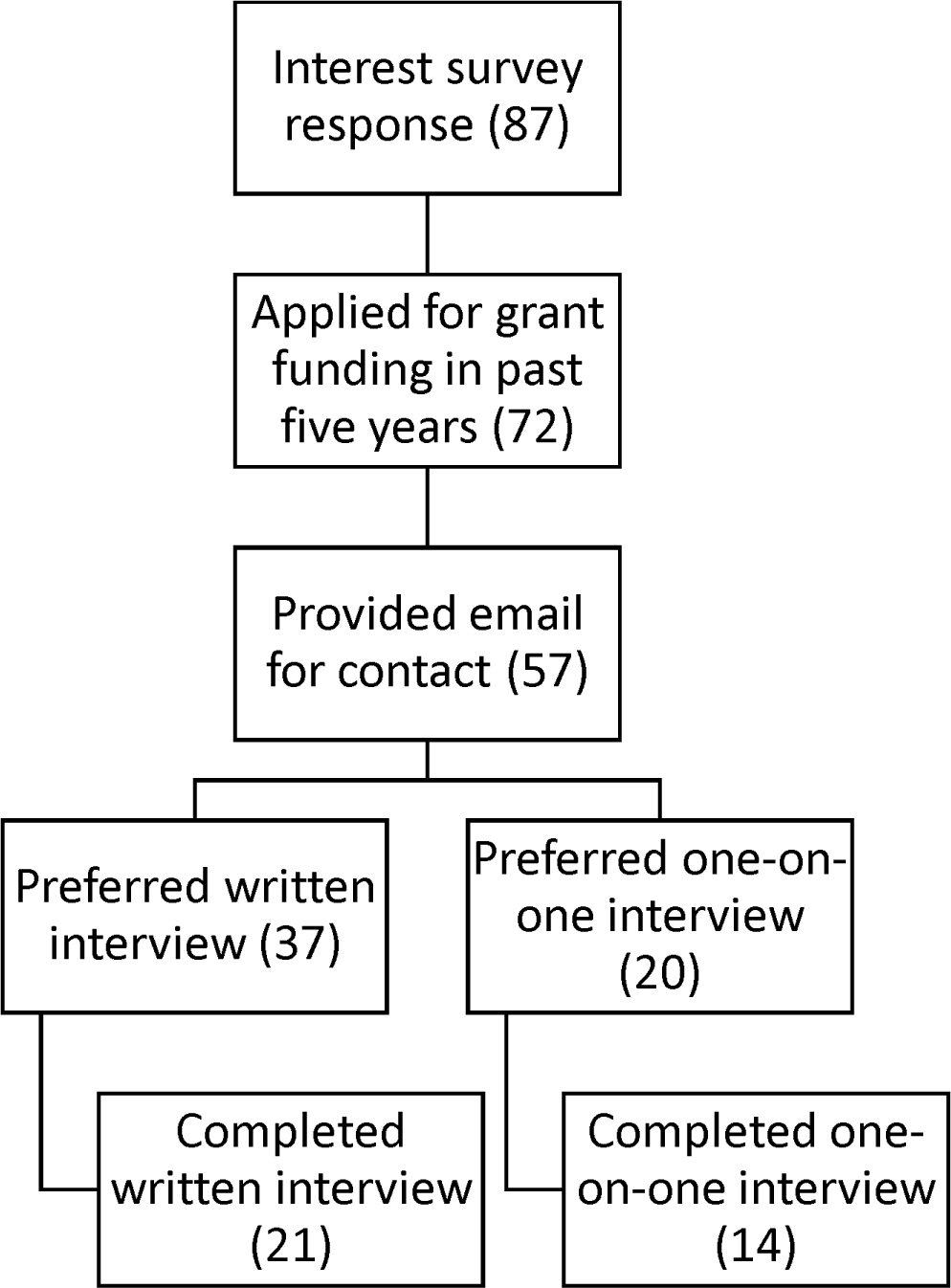
Participants’ selection and recruitment.

**Table 1.**
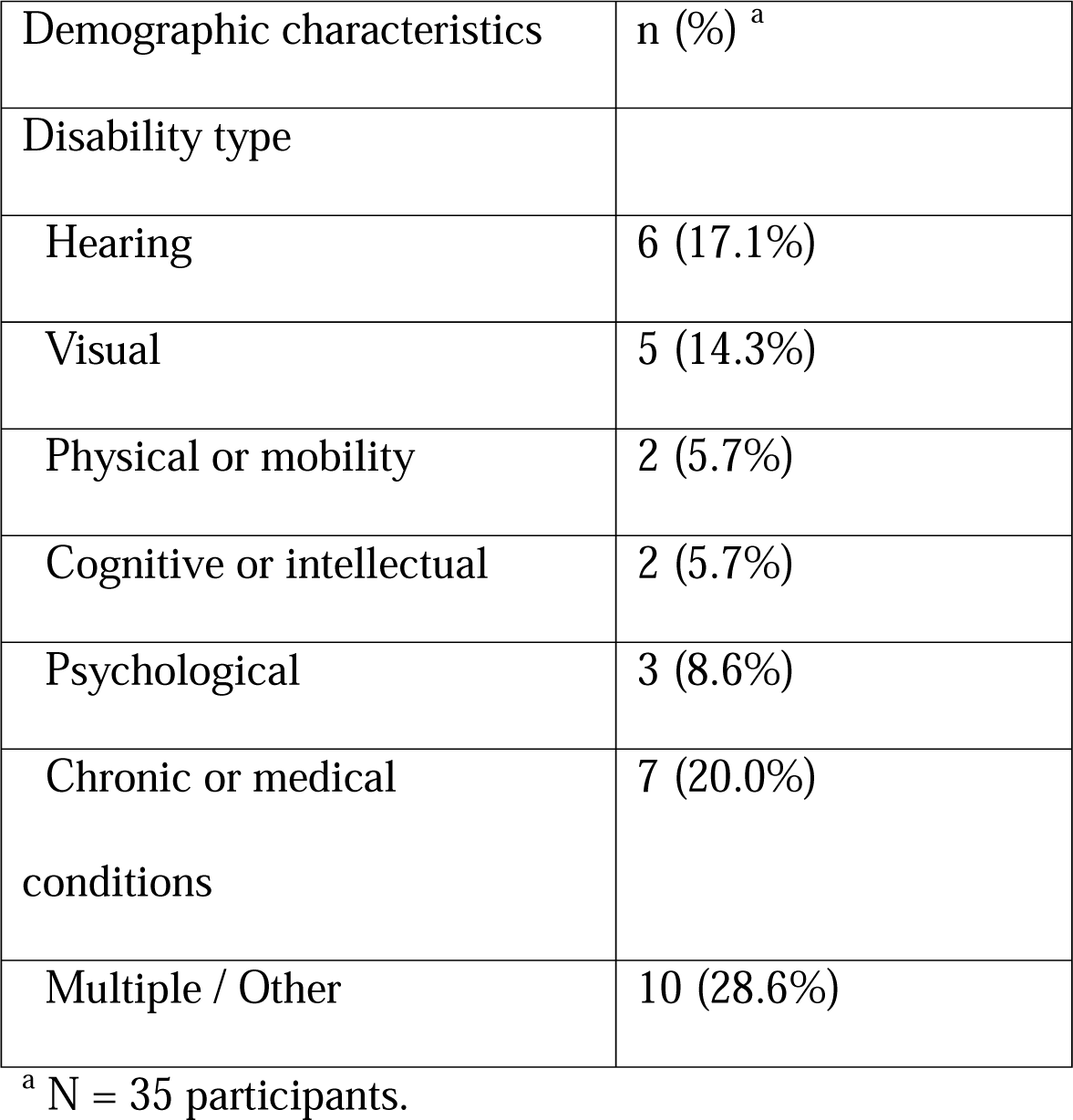
Participants’ disability types.

**Table 2.** Major subthemes described by participants.

| Subtheme |
| --- |
| Career trajectories |
| Identity, visibility, and disclosure |
| Academic relationships |
| Time management, sick leave, and work burden |
| Self-provision of accommodations |
| Accommodations provided by institutions |
| Accessibility obstacles |
| Bias, ableism, representation, and inclusion |
| COVID |

### Disability identity and bias in the workplace

Nine participants reported their disabilities as apparent to others, thirteen as non-apparent, and fourteen reported that the apparentness of their disability was somewhere in the middle, depending on whether they were appropriately accommodated or not. Accessibility aids such as wheelchairs, hearing aids, or tinted glasses were noted as a common way that people knew their disabilities were apparent to others. Having an apparent disability was beneficial for some participants, as it quickly demonstrated to others some basic access needs, whereas other participants stated it led to further discrimination. In some cases, participants did not identified as having a disability until decades later from when they first experienced limitations. This was often related to how they felt perceived by others, how they thought their access needs and experiences compared to others, and to when they received a diagnosis from a medical provider. Many people felt their disability identity was context-dependent. A participant said, “*It’s not the body that is disabling, it’s the intersection of a physical condition, deafness, blindness, whatever, and policies and practices that marginalize them*.” This illuminates why multiple participants were unsure whether they could identify as disabled if they had an impairment but did not experience as much stigma as they thought they needed in order to consider themselves disabled; one participant preferred instead to identify as someone with a chronic medical condition, and another reported that they felt much more comfortable identifying as disabled once they had an apparent disability. Others considered themselves disabled only when they noticed symptoms interfered with daily functioning. One individual said that after they began to identify as a person with a disability they looked back and realized that they have long made self-accommodations. Further, four participants with hearing loss discussed identifying as d/Deaf significantly before identifying as disabled, if they did at all, as “*a large part of the Deaf community rejects the label of disability*.”

After a diagnosis, several participants made changes to their research—seven specifically refocused onto disability-related topics. For some, this was entirely practical, as the physical environment or demands of their work were not accessible or accommodating. For others, this was a result of a new motivation towards disability research. The majority of participants gained new insight into their research, advocacy, and empathy for others as a result of their disability. This allowed researchers to connect more with their participants and also motivated them to strive for cross-disability access and equity.

Thirty-four participants described disclosing their disabilities or chronic conditions to at least some colleagues or students. Participants having apparent disabilities expressed that they did not have a choice in whether to disclose their disabilities. The primary reason participants limited their disclosure was fear of negative consequences such as being perceived as less capable by peers and leadership, and many described the process of disclosure as stressful or uncomfortable. One participant said, *“I have been told to hide disabilities from PIs [Principal Investigators] so they don’t treat me differently.”* Many participants avoided disclosing their disabilities during the hiring process, primarily because they had concerns they would be disqualified on the basis of assumptions made about their disabilities on the hiring side. Two individuals indicated concern that they would be seen as unobjective in their disability related research. One participant stated, *“Sometimes I think there’s a perception that researchers/academics with disabilities, especially those that disclose their disabilities, are only ‘advocates’, do less rigorous research, or are only there to disrupt. And while I do think of myself as an advocate, I also think it makes my research stronger and more impactful.”* The reasons people chose to disclose were much more varied, however, including: to better connect with disabled students or colleagues, to advocate more strongly for disability equity, for their direct safety or accommodation request, or to be seen as disabled rather than incapable.

Many participants commented on the unique complexity of disclosing their disability in job and grant application processes. Two participants described being very open about their disability in job interviews so they could ask interviewers direct questions about disability inclusion at the potential workplace. However, several participants were unwilling to disclose that they had a disability in a job interview for fear of discrimination. However, of these participants, some were willing to disclose their disability on a grant application, especially for applications based on disability research; this was because they hoped disclosure would demonstrate lived experience and bolster their application.

A total of 34 out of 35 participants raised the impact of ableism, stigma, and bias both at an institutional level and in professional relationships in academia. Participants commented on how leaving disability out of conversations around diversity, equity, and inclusion, constituted a form of exclusion at the organizational level. Networking was repeatedly noted as difficult, due to issues such as lack of masking to protect from disease such as COVID-19, hours that extend beyond 9 to 5, loud meeting areas, the exhaustion of travel, and more. At their own institutions, participants faced stigma from peers and leadership around their productivity, as one person stated, *“if someone takes leave for their own health, they are met with gossip, passive aggression and discrimination.”* Another participant said their superior suggested to them that their funding was acquired on the basis of their disability rather than merit. Many stated that bias prevented them from being able, or required them to work harder than others, to advance at a typical rate in their career, and in some cases led to people being pushed out of academia entirely. One person said, *“No one looks at my CV and recognizes that I was in incredible excruciating pain during this time and could not eat. Yet, despite that, I persevered. You don’t see that in my CV.”* One participant said they received lower teaching evaluations from students due to perceptions of their speech disability, and another reported, “*I was kicked out of my initial doctoral program when I was diagnosed with schizophrenia, and experienced significant disability-based discrimination in the doctoral program I graduated from”*.

These issues persisted at participants’ institutional disability service offices, where they found a lack of resources for faculty. One person stated that their experience was *“fully medicalized and overly dismissive of my actual … request.”* Several participants said that there is more attention to student access, with one person stating, *“over the course of my career in academia, it’s gotten progressively worse the further I get out from education.”* However, two participants found their institutions’ ADA attorneys helpful in navigating the accommodation process.

### Accommodations

Twenty-two participants stated that they had used accommodations in the workplace. Much of this required work in advance to set up an accommodating environment, such as organizing an accessible schedule for writing and meetings, apps to track tasks, preparing accessible transportation, and creating a checklist for access to review before each class. Others happened in the moment, such as notetaking and lip reading when captioning was not available. Participants self-accommodated with a wide range of assistive technology in their work, including portable microphones in the classroom, dual monitor systems, automatic captions, screen reading software, Braille display and notetakers, custom wheelchairs, white boards, and yellow tinted glasses.

Two people reported very positive, prompt provision of accommodations through official channels, though most had difficult experiences with delays or incorrect implementation. One individual shared, *“I didn’t expect grown adults to fight with me over something so simple. It took nothing from them, it took no time. I arranged everything for their schedule, and yet it was a fight, continually.”* Repeatedly, participants reported unsupportive leadership or administration. Another individual stated, *“No one is willing to change themselves for your disability. There’s accommodations, but they don’t transition into people giving you the accommodations.”* Participants said they experienced frequent neglect for their accommodation provision, whether the provisions were inconsistent, ignored, or forgotten, and that they were frustrated when left to manage their accommodations alone. One participant shared, *“I always thought that somehow there would be a system in place for helping that, but there isn’t. It’s all up to you and how well you navigate the system yourself. It’s very much you’re on your own.”*

One of the main reasons for delay were requirements for medical documentation of their disabilities. One participant describes the rigid process: *“The difficulty is a lot of medical situations don’t fit in the form because they require you to predict the future. There is no way to predict, essentially, a very unpredictable condition that knocks me out of practice for weeks at a time. There’s no way any accommodations process will do that, really. Everybody admits that.”* One participant described how getting documentation was difficult and used some discretionary account funds to pay for testing to find a diagnosis for their disability, *“There isn’t really support for figuring out, ‘You’ve got this, and this is what this means.’ I think it will make me better at my job if I could be like, ‘Yes. This is it. Therefore, these are the strategies that I do need to use’”*

Several participants, especially those with dynamic disabilities, stated they experienced accessibility difficulties in adhering to deadlines and other time-based expectations. One participant said, *“The whole problem with academia is it presumes that you’re able to put in a 60-hour week. Anybody who can’t put in a 60-hour week, whether it’s because they have little children, or they’re pregnant, or they’ve got a medical issue, or they’ve got a relapsing, remitting diagnosis, whatever, we suddenly discover that we have to find that time somewhere.”* Additionally, some participants described facing discrimination when requesting sick leave. This environment left some participants unsure of any accommodations that could assist them. Many participants found working from home to be a helpful tool, one made more readily available during the COVID-19 pandemic. Working from home was beneficial for time flexibility and productivity, as it allowed people to avoid physical pain, to turn on automatic captioning on video communication programs, to avoid sensory overload, and to take breaks or rest as needed. One person shared, *“The pandemic gave me the accommodations the institution wouldn’t and showed how easy it can be to integrate people with similar disabilities. I can manage my health condition on my time scale at my house while being productive, joining meetings, and mentoring.”*

### Advocacy

Respondents found an overall lack of disability inclusion and low representation of people with disabilities in academia. A participant shared, *“I think that there are initiatives to improve [disability inclusion], but I think that it is lacking.”* Participants found that they were underrepresented in diversity trainings and in academic positions, as one participant highlights: “*I regularly point out that a major weakness of our field is that enormous underrepresentation of direct experience among our ranks. It’s ridiculous that I’m regularly the only disabled person in a room, group, etc. focused on the study of people with disabilities.”* Another participant stated that these issues persist in publication and that “*publishers receive research about ableism and people with disabilities as very niche*,” stunting dissemination of the work.

Participants highlighted the importance of role models and mentors with disabilities, support systems, and community with other researchers with disabilities. The ability to share experiences and life trajectories not only constitutes a means of empowerment, but also helps in shaping career paths for the next generation of academics with disabilities. Several participants discussed mentoring students with disabilities, though this was typically unpaid labor. A participant stated, *“I use my experience with disability to provide a supportive and empathetic environment for students, primarily.”* Participants also felt supported by their students, as in the case of a participant whose students accommodated their visual disability by verbally describing tables and figures in group settings, *“For them I don’t often have to remind them or even ask them. Oftentimes they’ll ask me first. They’ll be like how can I do this best for you? What do you need? That is such a great interaction to have when somebody’s just aware and automatically wants to be inclusive.”* Another individual found community across institutions, and shared, *“The peer mentorship group with other scientists with hearing loss has been instrumental in pushing for accommodations at our annual research conference and providing social networking opportunities for us. We have even worked together to publish papers describing how scientific organizations can better support their scientists with disabilities.”* The absence of fellow disabled researchers in some instances led to difficulties in navigating disability-specific challenges in academia and a sense of isolation. One participant said, *“I’ve never had a mentor with a disability and generally find these challenges to be rather isolating to navigate alone”*.

In the face of extensive barriers, participants self-advocated for their access needs and understanding of their disability in academia. Three individuals discussed self-advocacy in advance or absence of negative experiences, such as for a blind professor who educated their students how to indicate they have a question or comment at the start of the semester. Self-advocacy also took place after access was denied, and participants highlighted that this was excessively time consuming of time intended for academics, and that this had to occur outside “official channels.” One person shared, *“I have to constantly advocate for myself… It’s been really hard to do that. It makes me really angry and frustrated. It takes up a lot of time, and it’s tiring.”* This participant, along with three others, referred to disability rights laws such as the Americans with Disabilities Act (ADA) when self-advocating to colleagues and leadership, when seeking accountability, and when protecting against unlawful intrusive questions about a disability.

Many participants engaged in disability advocacy that extended beyond themselves into their department, institution, community, and beyond. One person stated, *“I don’t want this just fixed for me. I want this fixed for everybody.”* Aims differed across participants, as some wanted disability inclusion in existing systems and recognition of disability as diversity, while others wanted systemic change that was “transformative” or created a “paradigm shift.” Respondents did this formally in leadership positions focused on diversity and access, as a part of their research, within disability advocacy groups, in public talks, as well as outside typical role requirements.

## Discussion

Our qualitative analysis of faculty with disabilities underscores the intricate, yet pervasive, obstacles faced by researchers with disabilities in academia. Our results reinforce previous research showing that faculty with disabilities encounter systematic barriers characterized by unaccommodating environments and structures traditionally rooted in bias and ableism.^13^ We further highlight how considerations around disability identity and disclosure are shaped by peers’ perceptions and fear of stigma and discrimination.

Brown and Leigh argue that “*ableism in academia is endemic*”,^15^ as the academic environment is often grounded on assumptions that cater to nondisabled people. Ableism impacts disabled researchers and faculty throughout their careers, including in educational, hiring, funding, and promotion phases. Many studies have shown how institutional policies reduce access to educational opportunities in science for young adults with disabilities,^7^ and how the pervasiveness of these barriers throughout the STEM educational pathway negatively impacts career choices, enrollment and retention in science-related degrees.^16^ Previous studies have shown that scientists with early-onset disabilities who work in academic institutions receive salaries that are $14,360 lower as compared to their nondisabled peers. Further, there has been a decline in National Institutes of Health (NIH) grant funding for disabled researchers, demonstrating lack of support for their work.^17^ This issue has received little attention, as previous research on faculty with disabilities has been excluded from higher education publications and each study is cited on average once a year.^12^

Furthermore, a study shows that disabled academics face discrimination and harassment at twice the rate of their nondisabled peers, a problem that compounds for people with multiple marginalized identities, such as LGBTQ people, Indigenous people, and women. Consequently, ableism impacts professional relationships between scientists with disabilities and students, peers, and leadership. Participants of this study reported experiences of social exclusion and marginalization when interacting with superior or peers, as also found in previous work.^18^ Negative attitudes and discrimination against disabled faculty hinder professional success, translate into barriers to securing and maintaining job positions, and lead to an attrition of disabled researchers in academia. Institutions’ views of disability as a medical construct limits the growth of scientists with disabilities and places the responsibility of access solely on the individual with a disability.^19,20^ Additionally, the frequent exclusion of disability from diversity discussions and initiatives not only perpetuates ableist institutional practices, but precludes the possibility of change towards more accommodating and inclusive higher education environments.^21^ There is a need to build stronger relationships between disabled faculty and their broader institutions through meaningful attitudinal and access changes as informed by people with disabilities’ lived experiences. ^22^ Intersectional approaches are critical to these changes, as disability is more prevalent among marginalized populations such as people of color, older adults, gender and sexual minorities, as well as people from lower educational attainment and income.^23,24^

Though Title 1 of the ADA requires employers to provide reasonable accommodation to employees with disabilities, inaccessibility and lack of disability inclusion are common to disabled faculty and students.^25^ In addition to mutual challenges, disabled postsecondary educators often lack accessible resources and must command authority in classrooms not designed for people with disabilities.^26^ Navigating accommodations is particularly challenging for faculty with disabilities as institutions often lack centralized disability resource centers, require lengthy processes for documenting a disability, and are sometimes unable to provide accommodations needed. At present, disability resource centers are often limited to students, leaving faculty to figure out accommodations on their own while dealing with unsupportive leadership or administrators. Participants stressed the experience of stigma when requesting and providing evidence for accommodations, and frustration due to the inflexibility of processes and reluctancy from institutions to providing accommodations, issues documented by previous studies.^27,28^ The Equal Employment Opportunity Commission (EEOC) regulates Title I of the ADA instead of the Department of Justice, with the former specializing on employment-related issues such as employment discrimination, and the latter having a broader mandate involving civil rights issues. While the existing regulatory framework allows employers to ask for reasonable medical documentation demonstrating an employee’s disability before providing a relevant reasonable accommodation, legal scholar Katherine MacFarlane criticizes this because it “*betrays the social model of disability on which the ADA rests and is inconsistent with legislative history and the EEOC’s own interactive process guidance*”.^29^

In light of the aforementioned barriers, participants frequently discussed how they engaged in advocacy to drive change, built community to help others navigate accommodations, and understood the importance of role models to foster the participation of people with disabilities in higher education. Due to low representation of intersectional minorities in academia, previous research shows the difficulties faced by individuals when they, beyond navigating obstacles of a system unaccommodating to their identities, constitute the only source of support and mentoring for those sharing the same identity.^30^ Our study aligns with previous research in highlighting how pivotal research and staff networks are as resources for peer-support and advocacy for scientists with disabilities.^31^ Furthermore, to truly advance a culture of inclusiveness, it is key that academic and research institutions are proactive about learning from and elevating not only the experiences of disabled scientists, but of the larger community of people with disabilities to ensure allyship and create welcoming spaces for disabled individuals.

Disability inclusion has recently gained attention in policy discussions in the United States. This includes the Biden administration’s executive orders to improve working conditions for people with disabilities in the federal workforce and to advance equity for underserved communities such as individuals with disabilities.^32,33^ Moreover, the National Institutes of Health recently recognized people with disabilities as a formal health disparities population,^34^ and organized a working group which developed recommendations to improve the inclusion of people with disabilities in research studies and the research workforce.^35^ These efforts have built momentum for further policy changes that support the inclusion and belonging of disabled people in research.

However, there remains a need for improved data collection on the experiences of faculty and researchers with disabilities, which are crucial for developing evidence-based approaches. This includes data that is longitudinal and disaggregated by racial and gender minority status, institution type, and career stage. The collection of these data could help identify institutions that have achieved measurable progress in improving the success and inclusion of disabled faculty and scientists. There is also need for harmonized databases across federal and state agencies collecting data on people with disabilities in educational institutions, and institutions should be incentivized to create centralized disability departments to expedite accommodations for scientists and faculty. A study using a novel university disability inclusion score highlighted that 60% of the top institutional recipients of NIH funding underperformed in inclusion and accessibility of their undergraduate programs.^5^ Moreover, given the importance of building networks in academia, strategies to make scientific meetings and conferences more accessible should be scaled-up to ensure increased participation of disabled scientists and facilitate the establishment of meaningful connections that could provide opportunities for mentoring, scientific collaborations, and peer-to-peer support.^36^

The results of this study should be interpreted while considering its limitations. As is the case for most qualitative studies, our sample was not representative of individuals by disability types, gender, race and ethnicity, or geographic location. Therefore, these results may not be generalizable across these groups and settings. This study primarily focused on individuals who were currently working as faculty or scientists, though future work should consider the experiences of individuals who ultimately left research or academic institutions for other sectors or unemployment.^6,37^ Questions from the screening questionnaire, particularly the one on type of disability, were open-ended. While this gave participants the opportunity to self-identify as having a certain disability, the data does not allow us to make granular comparisons as these categories differed from the ones used in previous studies. Strengths of this study should also be noted, such as the prioritization of accessibility in every step of the data collection process, and giving respondents flexibility to either engage in live or written interviews, which supports inclusion of participants across a diversity of disability groups. Additionally, our interviewers had lived experiences as people with disabilities, and this commonality has been shown to promote trust with participants. We also followed an approach of *centering disability*, which places disability as an integral part of the interview process that fosters flexibility and innovation.^38,39^

## Conclusion

This study contributes to our understanding of the experiences of faculty and scientists with disabilities in academia in the U.S. A lack of research and limited acknowledgement of obstacles impacting this community has held back changes, both at the institutional and government level, precluding the improvement of inclusion of disabled faculty and scientists. As Dunn points out, disability continues to be an afterthought at academic institutions, and while there has been progress in making higher education institutions more accessible and inclusive of people with disabilities, institutions are slow to remove physical and structural barriers.^40^ As a result, the overall experience of being disabled and working in academia is a tolling and complex undertaking that further deepens the already existing disparities for people with disabilities at higher education institutions, which perpetuates inequities stemming from reduced opportunities for participation in society.

Our results highlight that academic institutions have much work ahead to ensure faculty and researchers with disabilities are provided with equitable opportunities, resources, and supportive environments. Academic leadership must allocate resources to address and mitigate ableism, create more inclusive environments, and raise standards beyond the low bar of ADA compliance to promote equity, inclusion and belonging of disabled faculty.

## Data Availability

Data are not available as these were obtained from confidential interviews

---

**Supplementary Table 1.**
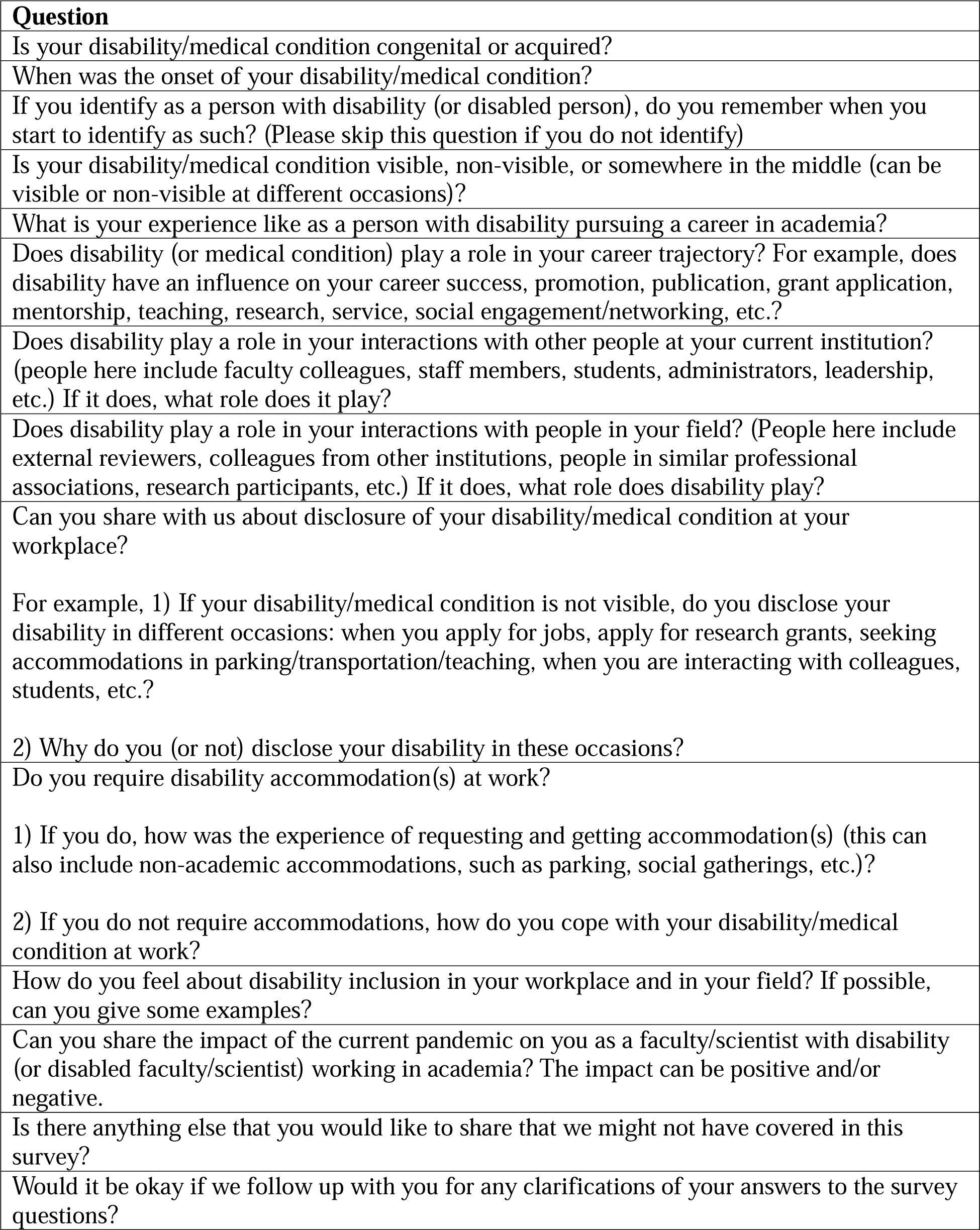
Interview Guide.

